# Evaluating Mid-Upper Arm Circumference Cut-Offs as a Screening Tool for Undernutrition in Pregnant Women: An Alternative to Body Mass Index

**DOI:** 10.64898/2026.02.25.26347073

**Authors:** Samanta Sabed, Israt Sharmin, Md Fuad Al Fidah, Ar-Rafi Khan, Fahmida Dil Farzana, Mehjabin Tishan Mahfuz, Gulshan Ara, Md Shabab Hossain, Tahmeed Ahmed, Mustafa Mahfuz

**Affiliations:** Nutrition Research Division, icddr,b, Mohakhali, Dhaka, Bangladesh; Office of the Executive Director, icddr,b, Mohakhali, Dhaka, Bangladesh

**Keywords:** Bangladesh, BMI, MUAC, Slum, Urban

## Abstract

**Background:** Undernutrition during pregnancy is a major public health issue and may lead to negative pregnancy outcomes. The body mass index (BMI), considered widely used as the reference method for assessing nutritional status due to its established population-based cut-off points; however, it can be misleading in pregnancy. This study aimed to validate the mid-upper arm circumference (MUAC) as a screening tool for identifying undernourished pregnant women (PW) in an urban slum, compared to BMI.

**Methods:** Data for this analysis were extracted from studies conducted in Bauniabadh, a slum located in Dhaka, Bangladesh. The final sample size was 375 PW aged 15-39 years with a gestational age of <14 weeks. The first recorded weight during enrolment was considered a proxy for pre-pregnancy weight. Participants were classified as undernourished or well-nourished accordingly. The BMI Z-scores for adolescents and the BMI categories for adult women were used to define undernutrition. Receiver operating characteristic (ROC) curve analysis was used to calculate the area under the curve (AUC), sensitivity, specificity, predictive values, and likelihood ratios. BMI-MUAC concordance was analyzed using McNemar’s test to determine optimal MUAC cut-offs.

**Results:** Among the candidate MUAC cut-off values, a threshold of <22.5 cm demonstrated high diagnostic accuracy, with 78.1% sensitivity, 92.9% specificity, an AUC of 85.5%, and a positive likelihood ratio (LR+) of 11.0. The alternative threshold of <23.0 cm showed higher sensitivity (84.4%) and AUC (86.9%) but lower specificity (89.4%) and LR+ (7.9). The difference in AUCs between the two thresholds was not statistically significant (p = 0.400). Using the <22.5 cm cut-off, 19.2% of pregnant women were classified as undernourished compared with 17.1% based on BMI. Concordance between MUAC <22.5 cm and BMI-defined undernutrition was satisfactory (p=0.182).

**Conclusions:** MUAC can be considered a simple and effective screening tool for identifying undernutrition in PW. Given its strong diagnostic accuracy, a threshold of <22.5 cm may be a practical alternative to BMI and considered for integration into nutritional programs for PW in Bangladesh.

## Introduction

Adequate maternal nutrition is essential for fetal growth and development. The prevalence of malnutrition in pregnant women (PW) in low- and middle-income countries (LMICs) remains a concerning public health issue. Undernutrition during pregnancy is disproportionately associated with increased risks of perinatal complications, preterm birth, low birth weight, maternal morbidity and mortality, especially in LMICs [1]. According to a UNICEF report, acute malnutrition among pregnant and breastfeeding women increased from 5.5 million in 2020 to 6.9 million in 2022, a 25% rise in 12 severely affected countries [2]. In addition, nearly half of the estimated 153.8 million underweight women of reproductive age worldwide reside in South Asia [3]. In South Asia and Africa, maternal malnutrition affects 25-33% of women [4, 5]. In Bangladesh, the prevalence of maternal underweight has markedly declined from 52% in 1996 to 19% in 2014 [3]. Unfortunately, the current prevalence still remains high compared to other LMICs [6]. To address this public health concern effectively, early detection of undernutrition in PW using simple, practical, low-cost, and measurable tools is essential.

Traditionally, body mass index (BMI) has served as a reference method for assessing nutritional status in PW. However, accurate calculation of BMI requires precise measurement of weight and height, which can be challenging in low-resource settings due to logistical constraints, limited equipment availability, and restricted mobility of participants [7]. Furthermore, BMI may be confounded by gestational weight gain, edema, or other health conditions, reducing its reliability during pregnancy [8]. Conversely, mid-upper arm circumference (MUAC) is a simple, cost-effective measure that does not require specialized equipment, making it suitable for large-scale community assessments [8]. Unlike BMI, MUAC remains relatively unaffected by pregnancy-related physical fluctuations, making it a more practical option for assessing nutritional status during this period [9]. MUAC is measured with a simple tape, enables quick detection of undernutrition and supports timely interventions. Although MUAC is a potential alternative, it has not yet replaced BMI as the standard metric due to the lack of universally accepted cut-off points for MUAC during pregnancy. Additionally, large-scale prospective validation across diverse populations remains limited [10].

Several studies have explored the relationship between MUAC and BMI [8, 11]. Research in India demonstrated MUAC’s potential as a valid measurement tool to identify undernutrition due to its ease of application [11]. Similarly, a study conducted in Bangladesh reported a strong correlation between MUAC and BMI in adults, proposing a MUAC cut-off of <24 cm for women as a simpler alternative to BMI [8]. However, that study did not focus on PW. The World Health Organization (WHO) recommends a MUAC cut-off between 21 and 23 cm among PW, emphasizing the importance of country- and context-specific thresholds for accuracy and suitability [12-14]. This study aimed to determine an optimal MUAC cut-off that aligns with the BMI-defined undernutrition threshold among PW in Bangladesh.

## Materials and Methods

### Data source description

We extracted data from two studies from two studies, namely “Nutrition for children, adolescent girls and pregnant women” (Nutri-CAP) and the “Maternal Environmental Enteric Dysfunction” (Maternal-EED) study. Both studies were conducted in Bauniabadh and adjacent slum areas of Dhaka, with an average population density of about 50,000 per square kilometer [15]. These communities are characterized by poverty, limited access to healthcare, and food insecurity. Participant enrollment started on 17 April 2022 and ended 15^th^ April 2024 for the Nutri-CAP study on the other hand for the Maternal-EED study enrollment started on 2 January 2023 and recruitment is ongoing. We included 375 PW aged 15-39 years for analysis in our study. PW younger than 15 years or beyond 14 weeks of pregnancy were excluded. BMI Z-scores were used for participants aged ≤19 years, while BMI categories were applied for others.

### Study procedure

According to the study objectives, datasets from both studies were merged, reshaped, and harmonized to ensure consistent variable definitions and measurement units. Anthropometric data on body weight, height, and MUAC were collected in both studies using standardized measurement procedures. Trained field workers performed all anthropometric measurements following standard techniques [16]. MUAC was measured on the non-dominant arm using an adult MUAC tape following the standard protocol [17]. Two consecutive measurements were taken for each participant, and the average value was recorded. To minimize observer bias, body weight and height were measured after MUAC assessment.

### Operational definitions

#### Undernourished pregnant mothers

We considered BMI Z-scores for participants aged ≤19 years [18] and BMI categories for adults to determine their nutritional status. According to WHO, adolescents are “thin” if their BMI Z-score is less than -2 SDs, corresponding to an adult BMI below 18.5 kg/m² [19]. Adolescents classified as thin were considered “undernourished”. Similarly, adult participants with a BMI of <18.5 kg/m^2^ were considered “undernourished”, following the WHO recommendations [20].

#### Independent variables

Independent variables in this analysis included age (in years), height (in cm), weight (in kg), MUAC (in cm), and duration of pregnancy (in weeks).

#### Pre-pregnancy BMI

In this analysis, only PW with gestational age <14 weeks at the time of enrolment was considered. Pre-pregnancy weight was not directly measured due to the study design. Therefore, the first recorded weight was used as a proxy for pre-pregnancy weight. Previous studies suggest that for PW from resource-poor contexts, weights recorded up to 16 weeks of pregnancy can serve as pre-pregnancy weight [21-24]. Evidence indicates that women in such settings gain minimal weight during this period. A study in South Benin also demonstrated strong concordance between measured pre-pregnancy weight and the weight up to 14 weeks of pregnancy [25]. This study adopted this approach.

### Statistical Analysis

The current study used STATA 19 (StataCorp LLC, College Station, Texas) for data processing, analysis, and graphical representation. Descriptive statistics included median and interquartile range (IQR), or frequency and percentage. We assessed the predictive accuracy of MUAC for identifying undernourished PW by computing the area under the receiver operating characteristic curve (AUC). The “roccomp” command in STATA was used to compare AUC values of different MUAC cut-offs. We assessed the effectiveness of MUAC cut-off points compared to BMI categorization using AUC, sensitivity, specificity, positive predictive value (PPV), negative predictive value (NPV), and likelihood ratios. The candidate thresholds started from 21 cm up to 23 cm at 0.5 cm intervals. Several international organizations recommend this range [24]. As per their guideline, the MUAC cut-off with the highest sensitivity at or above 70% specificity was considered as a candidate for the optimum cut-off. The AUC represents overall discriminative ability, interpreted as: excellent (0.9-1), good (0.8-0.9), fair (0.7-0.8), poor (0.6-0.7), and fail (0.5-0.6) [30]. An AUC of 0.5 indicates no discriminative power beyond chance. The AUC values reported by the candidate thresholds were compared using the “roccomp” command from STATA. The exact McNemar’s test was applied to examine concordance between the optimal MUAC threshold (<22.5 cm) and BMI classification. The results indicate concordance between BMI and MUAC in identifying nutritional status among pregnant women (p=0.182).

We also considered other parameters like the positive likelihood ratio (LR+) and the negative likelihood ratio (LR-). It has been suggested that an LR+ of 10 is indicative evidence to rule in the diagnosis, i.e., considering the condition to be present [26]. In this context, the LR+ indicates the likelihood that a true undernourished participant (identified using BMI) is going to be classified as such using the MUAC cut-off. Conversely, the negative likelihood ratio (LR−) expresses the likelihood that MUAC accurately identifies participants without undernutrition as such. However, given the nature and objective of the study, the LR+ was given more importance during interpretation. All statistical tests were two-tailed, and a p < 0.05 was considered significant.

## Results

The study analyzed 375 participants whose age was 15-39 years. The process of participant selection is shown in Figure 1.

**Figure 1.**
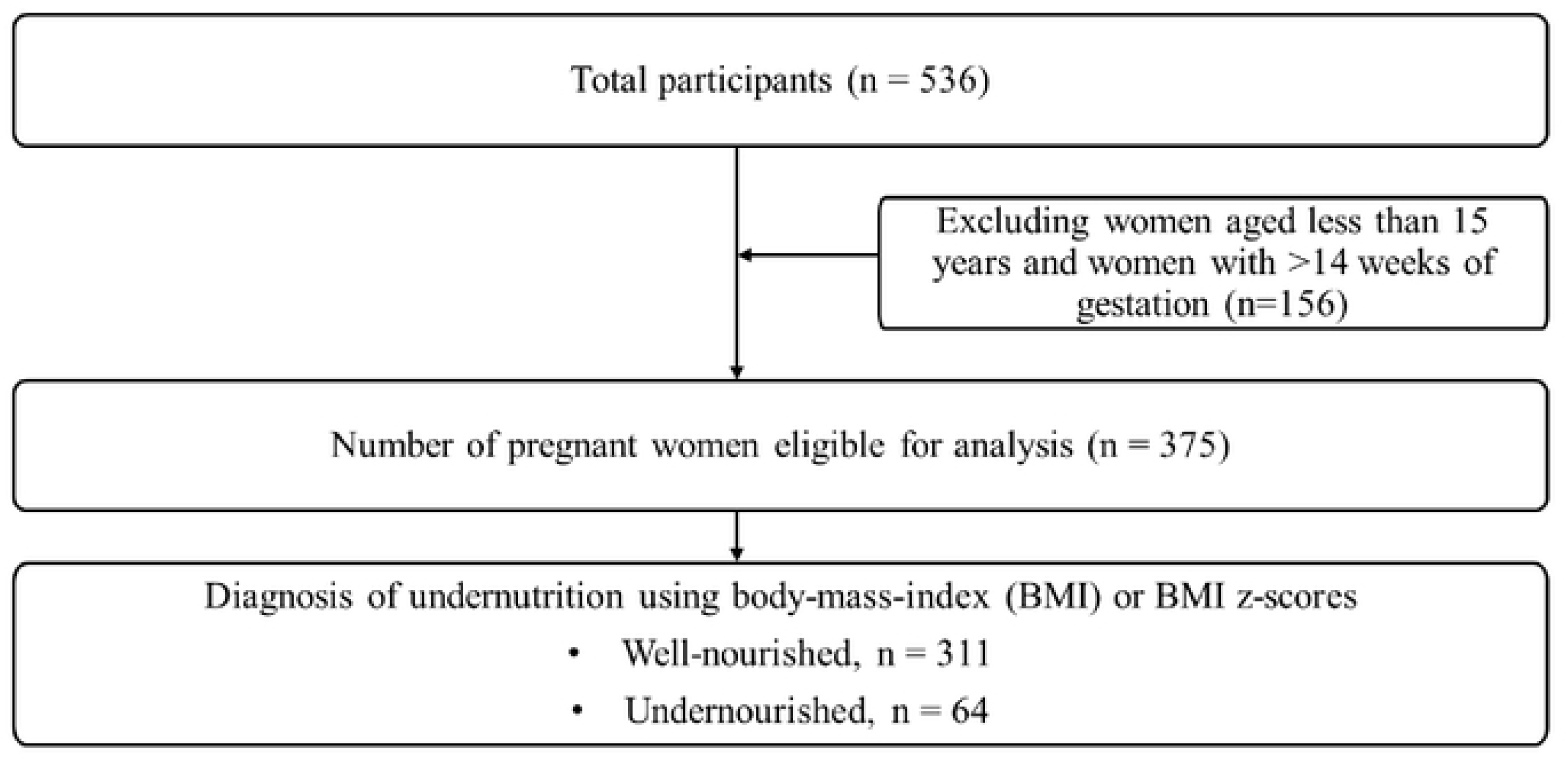
Study flowchart.

Baseline characteristics of participants are presented in Table 1. The median (IQR) age, MUAC, and duration of gestation were 22.4 (19.6-27.3) years, 26.5 (23.2-29.6) cm, and 12.0 (12.0-13.0) weeks, respectively.

**Table 1:**
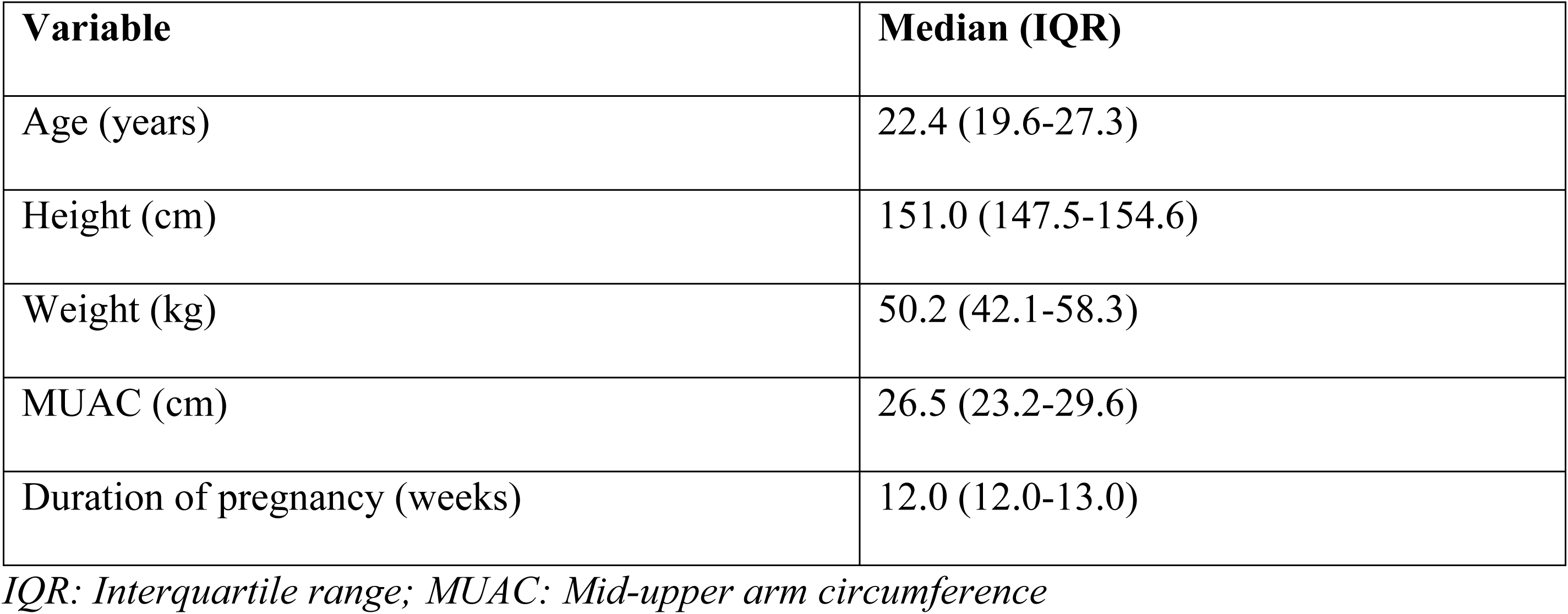
Characteristics of study participants.

| Variable | Median (IQR) |
| --- | --- |
| Age (years) | 22.4 (19.6-27.3) |
| Height (cm) | 151.0 (147.5-154.6) |
| Weight (kg) | 50.2 (42.1-58.3) |
| MUAC (cm) | 26.5 (23.2-29.6) |
| Duration of pregnancy (weeks) | 12.0 (12.0-13.0) |

*IQR: Interquartile range; MUAC: Mid-upper arm circumference*

Figure 2 shows that 17.1% of PW were undernourished according to BMI.

**Figure 2.**
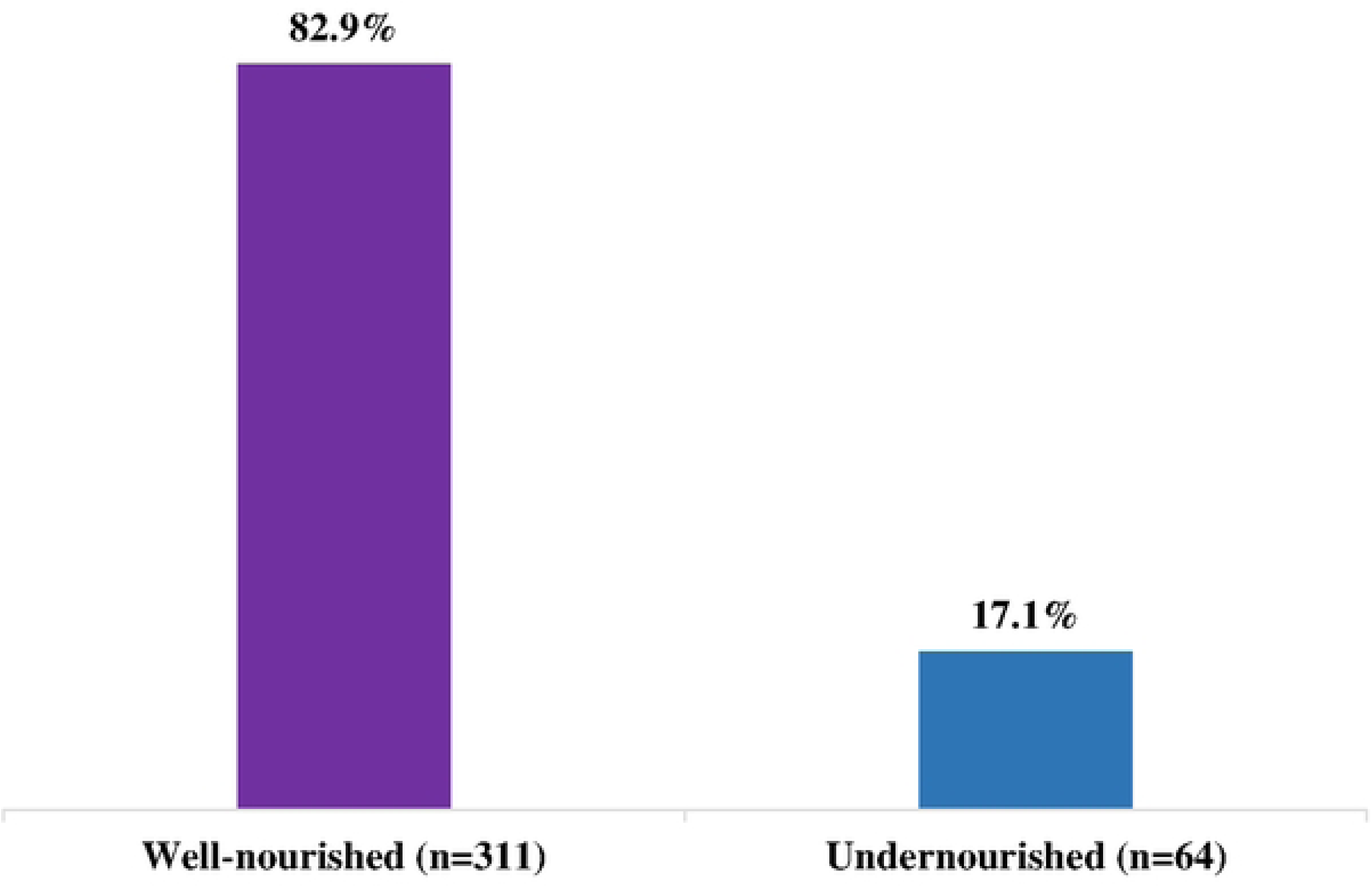
Nutritional status of pregnant women.

Performance metrics of various MUAC cut-offs are presented in Table 2. In general, as the cut-off points increase, the sensitivity increases but the specificity decreases. The optimal MUAC cut-off was <22.5 cm, with high specificity (92.9%), good sensitivity (78.1%), AUC (85.5%), PPV (69.4%) and LR+ (11.0). The alternative cut-off of <23 cm also showed high specificity (89.4%), sensitivity (84.4%), AUC (86.9%); however, its LR+ and the PPV were lower (8.0 and 62.1%, respectively).

**Table 2.** Performance of different MUAC cut-off values for identifying undernourished PW.

| **Cut-off point** | **Sensitivity** | **Specificity** | **LR+** | **LR-** | **PPV** | **NPV** | **AUC** |
| --- | --- | --- | --- | --- | --- | --- | --- |
| **<21 cm** | 43.8 | 99.0 | 45.35 | 0.57 | 90.3 | 89.5 | 71.4 |
| **<21.5 cm** | 51.6 | 96.8 | 16.04 | 0.50 | 76.7 | 90.7 | 74.2 |
| **<22 cm** | 59.4 | 95.5 | 13.19 | 0.43 | 73.1 | 92.0 | 77.4 |
| **<22.5 cm** | 78.1 | 92.9 | 11.04 | 0.24 | 69.4 | 95.4 | 85.5 |
| **<23 cm** | 84.4 | 89.4 | 7.95 | 9.17 | 62.1 | 96.5 | 86.9 |

*LR+: Positive likelihood ratio; LR-: Negative likelihood ratio; PPV: Positive predictive value;*
*NPV: Negative predictive value; AUC: Area under the receiver operating curve*

(Figure 3A) shows the receiver operating characteristic (ROC) curve comparing MUAC <22.5 cm with BMI-defined undernutrition among PW. The AUC was 85.5% for MUAC <22.5 cm and 86.9% for MUAC <23.0 cm, indicating comparable discriminatory ability (p=0.400) (Figure 3A). McNemar’s test reported a good concordance between MUAC <22.5 cm and BMI-defined undernutrition (p=0.182) (Figure 3B). We used a predetermined criteria to identify the MUAC threshold that best aligns with BMI classification for analysis, excluding cut-offs that showed lower consistency. This approach identified MUAC <22.5 cm as the most appropriate threshold for detecting low BMI (Figure 3C).

**Figure 3A:**
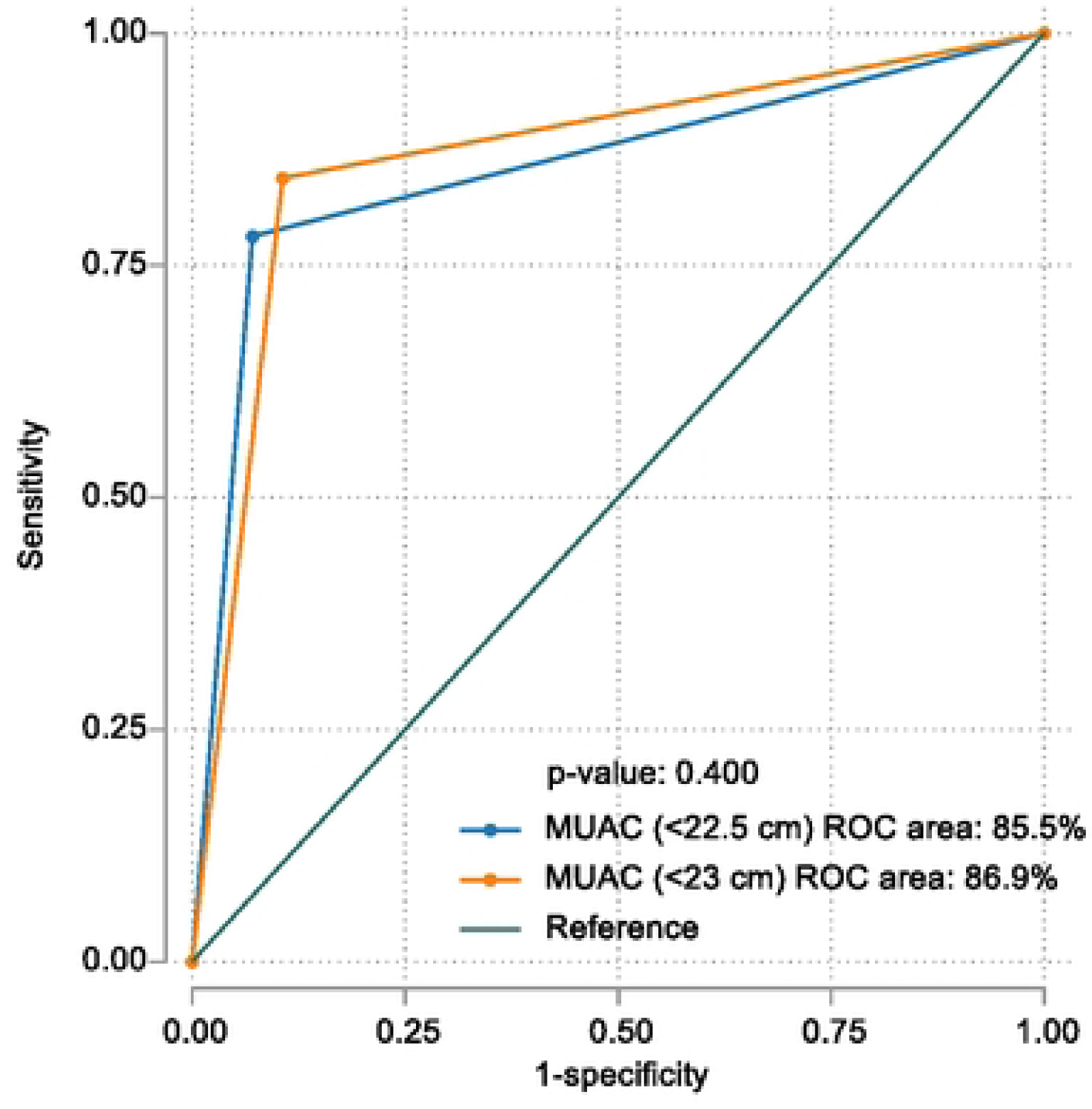
Comparison of AUC (<22.5 cm and <23 cm)

**Figure 3B:**
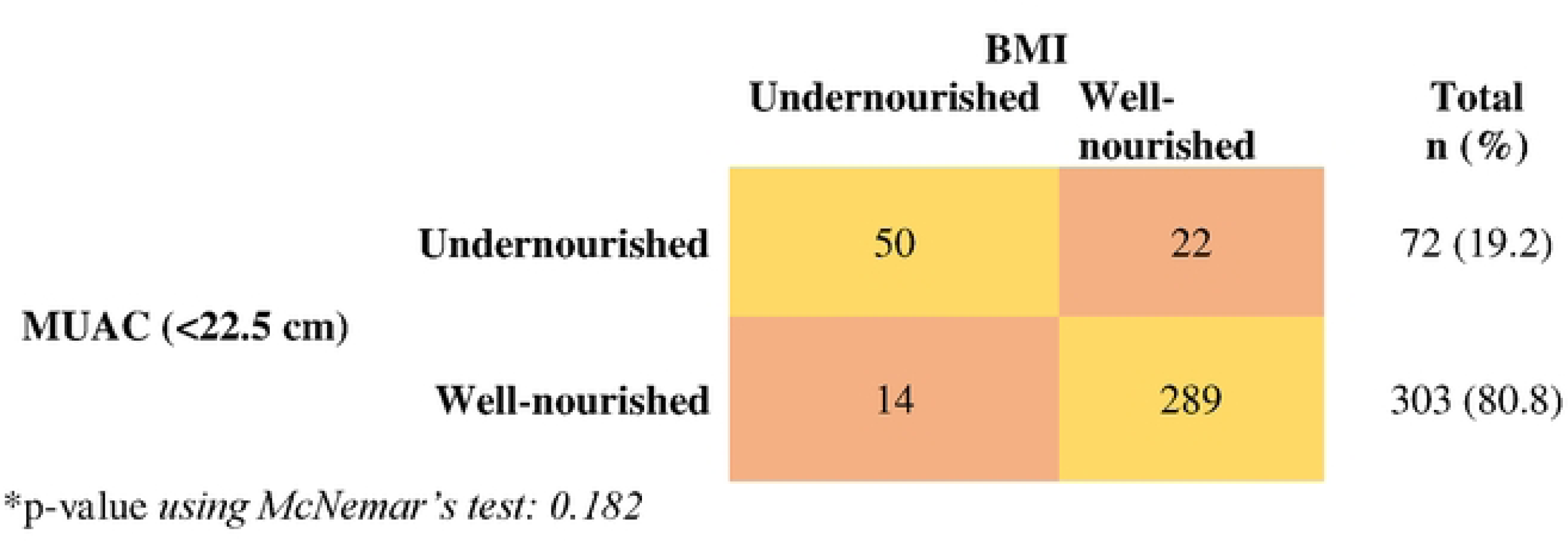
Concordance between BMI and MUAC categories.

**Figure 3C:**
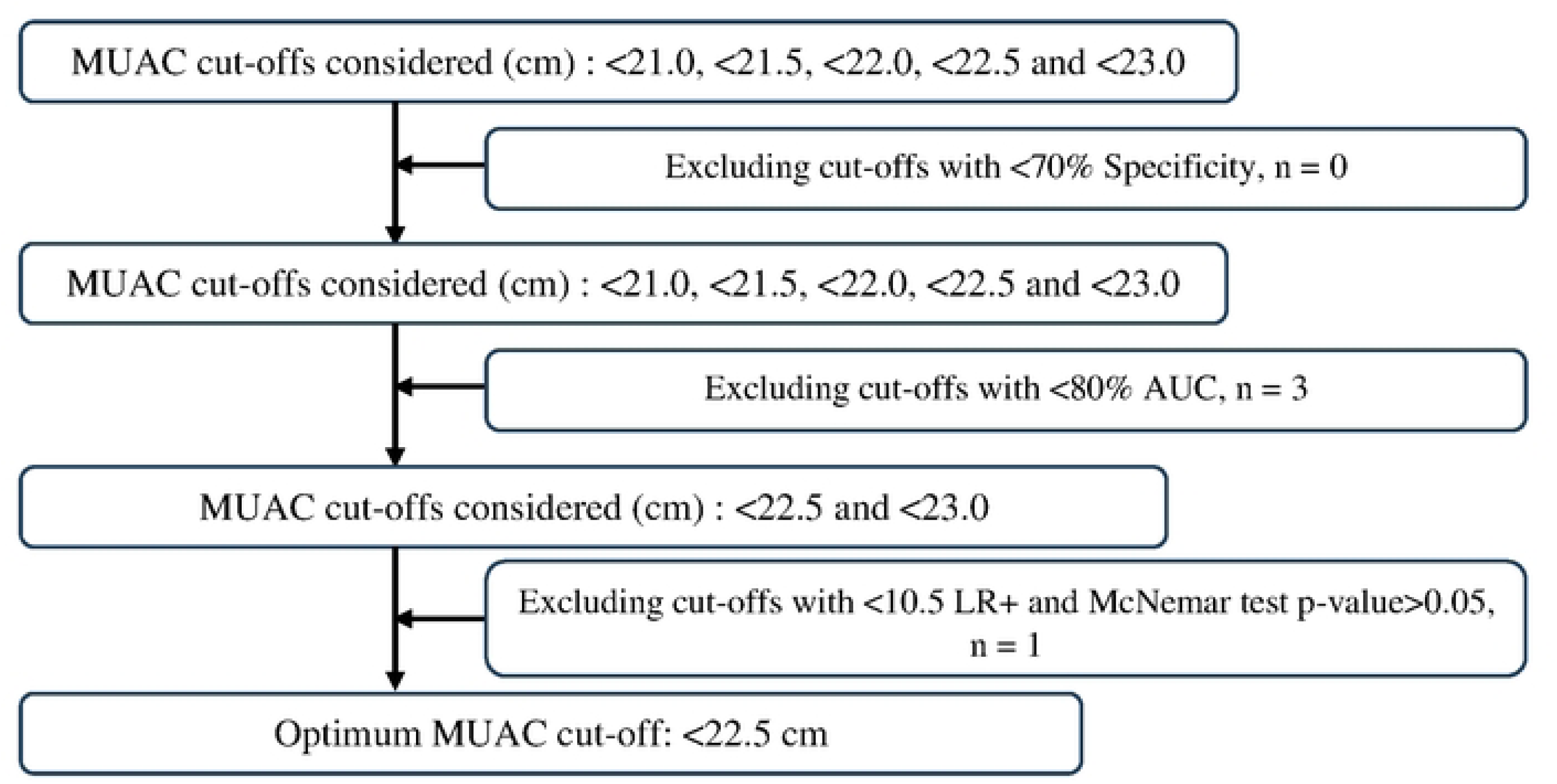
Steps of criterion-based evaluation for determining the optimum MUAC cut-off point Discussion.

Accurate BMI measurement requires technical skill and can be difficult in resource-limited settings or among individuals with health conditions. For example, BMI cannot be calculated for participants who are unable to stand [8]. In contrast, MUAC is easier and only needs a measuring tape and minimal training [8]. Moreover, it has a clear advantage over BMI during pregnancy.

BMI is less reliable during pregnancy because it is influenced by gestational weight gain and other physiological factors [8]. Approximately one-fifth of the PW were “undernourished” according to their BMI or BMI Z-scores. The median MUAC was 26.2 cm. Our analysis showed that setting the threshold at <22.5 cm yielded high specificity and sensitivity. Additionally, the LR+ was 11.0, indicating strong diagnostic performance for identifying undernourished pregnant women. The guidelines suggest that the highest sensitivity at or above a predefined specificity should be considered for screening purposes [13]. We considered a positive likelihood ratio (LR+) greater than 10 as the optimal diagnostic threshold [26]. Previous literature suggests that an LR+ above 10 provides strong evidence to “rule in” a diagnosis [26]. Although the MUAC cut-off of <23.0 cm demonstrated higher sensitivity than <22.5 cm, its specificity was lower (89.4% vs. 92.9%). The AUC difference between the two thresholds was not statistically significant, and the PPV for <23.0 cm was lower. Additionally, the LR+ for <23.0 cm (7.9) did not meet the desired benchmark of 10.

In considering the classification of nutritional status, we compared BMI and MUAC based classifications using McNemar’s test. The difference between the two methods was not statistically significant (p = 0.182), indicating satisfactory agreement in identifying undernourished pregnant women. The <22.5 cm cut-off was more specific and is recommended by this study, as it misclassifies fewer well-nourished individuals as undernourished. This criterion is critical for ensuring that nutritional interventions reach at-risk pregnant women in resource-limited settings like Bangladesh.

Several studies have supported the use of MUAC as a simple and practical tool for detecting malnutrition among pregnant women, particularly in resource-limited settings [7, 12-14, 27, 28]. The WHO and Sphere guidelines classify undernutrition at MUAC <21 cm and <23 cm, whereas FANTA recommends country-specific guidelines based on sensitivity and specificity [12-14].

For instance, research in eastern Ethiopia identified malnutrition using a MUAC threshold of <22 cm, highlighting its usefulness for assessing nutritional status during pregnancy [29]. Similarly, studies in Sudan have proposed that a MUAC cut-off point of 22.5 cm effectively identifies undernourished women [27]. Consequently, MUAC has been widely integrated into prenatal care as a simple anthropometric indicator for screening maternal undernutrition, particularly in low-resource settings [30]. A study in Spain involving 1,373 participants from diverse age groups demonstrated that a MUAC threshold of ≤22.5 cm correlated strongly with BMI, confirming its relevance across varied populations [31].

This study analyzed data from 375 pregnant women in a low-resource setting in Bangladesh and identified a MUAC cut-off of <22.5 cm as an effective threshold for detecting undernutrition among PW. Standardized measurement procedures and appropriate statistical tests were performed to strengthen the reliability of the findings. However, several limitations should be noted.

First, the cross-sectional design of the study limits the assessment of temporality. Second, BMI was used as a reference standard, which may not be ideal during pregnancy because of weight gain, gestational age and edema. Future research should consider the comparison of MUAC with other validated body composition methods, such as bioelectric impedance analysis (BIA).

Finally, all participants in this analysis came from a low-income urban community, which limits national representativeness. Therefore, the findings may not be generalizable to the broader population.

## Conclusion

The findings suggest that MUAC is a simple, practical, and reliable tool for identifying undernutrition among pregnant women, especially in low-resource settings. Based on the findings, a MUAC cut-off of <22.5 cm offers strong diagnostic accuracy and a balanced trade-off between sensitivity and specificity. This makes MUAC a useful alternative to BMI, which may be difficult to measure in some contexts. Because MUAC is easy to measure and requires little training and minimal equipment, it could be integrated in maternal health programs in Bangladesh. However, future validation studies should include nationally representative samples to ensure the generalizability of these results.

## Author Contributions

Conceptualization: TA, SS; Data curation: SS, ARK, IS; GA; Formal analysis: ARK, MFAF, SS, IS, TM; Methodology: MFAF, SS, ARK, IS, MM, TM, MSH; Writing original draft: SS, MFAF; ARK, MSH, FDF; Writing – review & editing: GA, MM, TA, MSH, FDF; Supervision: MM; Guarantor: SS

## Acknowledgements

Data used in this study were extracted from the “Nutrition for Children, Adolescent Girls and Pregnant Women” (Nutri-CAP) and the “Maternal Environmental Enteric Dysfunction” (Maternal-EED) studies.

Financial Support: The Nutri-CAP study was funded by the Department of Foreign Affairs, Trade and Development (DFATD), through Advancing Sexual and Reproductive Health and Rights (AdSEARCH), Grant number: SGDE-EDRMS #9926532, Project P007358. The Maternal- EED study was funded by The Bill and Melinda Gates Foundation (BMGF), Grant number: 02273, Project 22117. icddr,b is also grateful to the Government of Bangladesh and Canada for providing core support.

## Availability of data

Data pertaining to this manuscript can be obtained upon request. Researchers who meet the criteria for access to confidential data are encouraged to contact Shiblee Sayeed at the Research Administration of the icddr,b (http://www.icddrb.org/).

## Ethical Standards Disclosure

All procedures involving research, consent of study participants, data analysis, and publication were approved by the Institutional Review Board (IRB) of the International Centre for Diarrhoeal Disease Research, Bangladesh (icddr,b), which comprises the Research Review Committee (RRC) and Ethical Review Committee (ERC) (ClinicalTrials.gov, identifier-NCT05311436).

